# IMPACT OF GLUCOCORTICOID TREATMENT IN SARS-COV-2 INFECTION MORTALITY: A RETROSPECTIVE CONTROLLED COHORT STUDY

**DOI:** 10.1101/2020.05.22.20110544

**Authors:** Ana Fernández Cruz, Belén Ruiz-Antorán, Ana Muñoz Gómez, Aránzazu Sancho López, Patricia Mills Sánchez, Gustavo Adolfo Centeno Soto, Silvia Blanco Alonso, Laura Javaloyes Garachana, Amy Galán Gómez, Ángela Valencia Alijo, Javier Gómez Irusta, Concepción Payares-Herrera, Ignacio Morrás Torre, Enrique Sánchez Chica, Laura Delgado Téllez de Cepeda, Alejandro Callejas Díaz, Antonio Ramos Martínez, Elena Múñez Rubio, Cristina Avendaño-Solá, on behalf of Puerta de Hierro COVID-19 Study Group

**Author notes:** AFC and BRA contributed equally to this article. **Author for correspondence:** Dr. Ana Fernández Cruz, Infectious Diseases Unit, Internal Medicine Department, Hospital Universitario Puerta de Hierro-Majadahonda, Madrid, Spain. **Alternative corresponding author:** Dr. Belén Ruiz-Antorán, Clinical Pharmacology Department, Hospital Universitario Puerta de Hierro-Majadahonda, Madrid, Spain.

## Abstract

**Objective:** We aim to determine the impact of steroid use in COVID-19 pneumonia in-hospital mortality.

**Design:** We performed a single-centre retrospective cohort study.

**Setting:** A University hospital in Madrid, Spain, during March 2020.

**Participants:** Patients admitted with SARS-CoV-2 pneumonia.

**Exposures:** Patients treated with steroids were compared to patients not treated with steroids. A propensity-score for steroid treatment was developed. Different steroid regimens were also compared, and adjusted with a second propensity score.

**Main Outcomes and Measures:** To determine the role of steroids in in-hospital mortality, univariable and multivariable analyses were performed, and adjusted including the propensity score as a covariate. Survival times were compared using a log-rank test.

**Results:** During the study period, 463 out of 848 hospitalized patients with COVID19 pneumonia fulfilled inclusion criteria. Among them, 396 (46.7%) consecutive patients were treated with steroids and 67 patients were assigned to the control cohort. Global mortality was 15.1%. Median time to steroid treatment from symptom onset was 10 days (IQR 8 to13). In-hospital mortality was lower in patients treated with steroids than in controls (13.9% [55/396] versus 23.9% [16/67], OR 0.51 [0.27 to 0.96], p= 0.044). Steroid treatment reduced mortality by 41.8% relative to no steroid treatment (RRR 0,42 [0.048 to 0.65). Initial treatment with 1 mg/kg/day of methylprednisolone (or equivalent) versus steroid pulses was not associated with in-hospital mortality (13.5% [42/310] versus 15.1% [13/86], OR 0.880 [0.449-1.726], p=0.710).

**Conclusions:** Our results show that survival of patients with SARS-CoV2 pneumonia is higher in patients treated with glucocorticoids than in those not treated. In-hospital mortality was not different between initial regimens of 1 mg/kg/day of methylprednisolone or equivalent and glucocorticoid pulses. These results support the use of glucocorticoids in SARS-CoV2 infection.

**Summary:** We investigated in-hospital mortality of patients with SARS-CoV-2 pneumonia in a large series of patients treated with steroids compared to controls, and adjusted using a propensity score. Our results show a beneficial impact of steroid treatment in SARS-CoV-2 pneumonia.

## INTRODUCTION

Infection with the new coronavirus, SARS-CoV-2, presents mainly with respiratory involvement. Clinical presentation consists of a first viremic phase 7-10 days long, followed in some cases by a second phase of clinical manifestations driven by lung and systemic inflammation ^1^.

During the initial viremic phase, antiviral drugs are recommended, especially in cases with pneumonia. Around 80% of cases will resolve after this first phase. However, another 20% will evolve to a severe pneumonitis, followed by acute respiratory distress syndrome (ARDS).

In that second phase, an increase in acute phase reactants and macrophage activation markers has been identified. Poor outcomes have been associated with high IL6 levels, leading to the recommendation of treatment with IL6 antagonists.

Scarcity of anti-inflammatory targeted therapies such as tocilizumab during the initial period of SARS-CoV2 pandemic has driven the use of glucocorticoids in these patients, particularly in the more severe cases lacking other alternatives as a last-resort, despite the recommendation against it. Based on studies performed during the prior SARS-CoV, MERS-CoV, and H1N1 influenza epidemics, glucocorticoids were advised against in COVID-19 by the WHO, owing to a possible deleterious effect of prolongation of viral excretion and increased adverse events ^2^. The available studies have important methodologic limitations and, of note, glucocorticosteroids (from now on, steroids) were usually administered early after symptom onset (4 days)^3^. Nevertheless, in the current pandemic, the Chinese National Commission recommended methylprednisolone 1-2 mg/kg/day during 3-5 days in cases with respiratory failure ^4^ and several studies suggest a possible beneficial effect of steroids administered in the inflammatory phase of the disease, in patients with ARDS^5^. In this respect, the use of steroids as adjuvant therapy in moderate to severe ARDS is accepted in early stages at a dosage of 1 mg/kg/day of methylprednisolone in intubated patients^6^. In severe and rapidly progressive ARDS, methylprednisolone appears to improve symptoms and pulmonary damage, but does not increase survival^7^. Nevertheless, this inhibition of the inflammatory storm may allow to gain time to control the infection and prevent secondary multi-organ failure and shock ^8^ Recently, early administration of dexamethasone has shown a survival advantage in established moderate-to-severe ARDS^9^. Moreover, the combined effect of steroids with other anti-inflammatory therapies used concomitantly is still to be determined, namely in those cases in which the targeted therapy needs some time to achieve a response ^8^.

However, there is a lack of good quality evidence to support steroid use, and there is uncertainty not only about the overall effects, but also about the most appropriate drug, dose and timing. It is unknown whether the appropriate steroid dose might be the same in different moments of the disease, and which is the therapeutic ceiling.

While awaiting for results from ongoing clinical trials^10^, we consider that an analysis of actual clinical practice is needed to guide the recommendations. We hypothesize that steroid use can improve the mortality of patients with COVID-19 pneumonia, and we performed a retrospective analysis of our experience in a University Hospital in Madrid in March 2020.

## PATIENTS AND METHODS

### 1. Design, study period and subjects

This single-center retrospective cohort study included patients admitted to Hospital Puerta de Hierro-Majadahonda between March 4th, 2020, and April 7th, 2020. Our institution is a 613-bed tertiary teaching hospital in Madrid, Spain.

Adult patients diagnosed with COVID-19 pneumonia according to WHO interim guidance, and complicated with ARDS and/or an hyperinflammatory syndrome, where included. Of them, patients who received corticosteroid therapy according to clinical practice were assigned to the steroid cohort, whilst patients who did not were assigned to the control cohort.

### 2. Data collection

Epidemiological, clinical, laboratory and radiologic data, including concomitant COVID-19 treatments, were extracted from electronic medical records (SELENE System, Cerner Iberia, S.L.U., Madrid [Spain]) using a standardized data collection form. All data were included by a primary reviewer and subsequently checked by two senior physicians.

### 3. Laboratory procedures

Routine blood examinations included a complete blood count, coagulation profile, serum biochemical tests (including lactate dehydrogenase), C reactive protein, D Dimer, interleukin-6 (IL-6), and serum ferritin. Chest radiographs or CT scans were also done for all inpatients.

### 4. Definition of the outcome

The main outcome variable was in-hospital mortality. The outcome of patients treated with steroids was compared to that of those who did not receive steroids.

### 5. Definition of the exposure

Exposure to corticosteroids was defined as the use of intravenous steroids at any time during the hospital admission.

Patients with steroid treatment were designated as the “treatment cohort”, and those who did not receive them as “control cohort”.

The decision to prescribe steroids was at the discretion of the treating physician as the use of corticosteroids was not included in the COVID local protocol at the time of the study. Details of corticosteroid use (including the timing of initiation, dosing, and type of medications) were recorded. Likewise, the choice of COVID treatments other than corticosteroids was at the discretion of the treating physician, although based on national and local recommendations for COVID-19 management.

In the treatment cohort, the first day of administration was considered as the index date (day 0). In the control cohort, the index date was selected as the date at which the patient fulfilled ARDS criteria or presented any inflammation-related parameter (D-dimer, CRP, ferritin, IL6, lymphocyte counts) level over the limits of normal range. The diagnosis and grading of ARDS was determined according to modified Berlin criteria^11^.

For the main analysis, we generated a variable with the following mutually exclusive categories: “non-use of steroids drug” (control cohort) and “use of steroids drug” (treatment cohort). Subsequently, we disaggregated the category “use of steroids drug” into two different subgroups: 1 mg/kg/day methylprednisolone or equivalent, and steroid pulse. When a patient received different corticosteroid regimens during hospitalization, the first prescribed regimen was considered for the analysis.

### 6. Statistical analysis

Quantitative variables were expressed as means and standard deviations (SD) and/or medians and interquartile ranges, and qualitative variables as frequencies and percentages. The association of comorbidities among the treatment and control cohorts with mortality was assessed through univariable conditional logistic regression to compute crude odds ratios (ORs) and their 95% confidence intervals (CIs). Survival times were estimated using the Kaplan–Meier method, differences between the cohorts were compared using a log-rank test. The Mann-Whitney U test, χ^2^ test, or Fisher’s exact test were used to compare differences between survivors and non-survivors, where appropriate. To explore risk factors associated with in-hospital death, univariable and multivariable logistic regression models were used. Variables with a p < 0.05 in univariable models were selected into the multivariable.

To reduce the effect of corticosteroid treatment selection bias and potential confounding, we adjusted for differences in baseline characteristics by a propensity score, which predicts the patient’s probability of being treated with steroids regardless of confounding factors, using multivariable logistic regression. Potential confounders considered in propensity score matching (PSM) analysis were those variables included in the final model by means of step-wise backward elimination procedures. The effect of corticosteroid treatment on clinical outcome was analyzed by a multivariable logistic regression, adjusted for major variables associated with mortality; the individual propensity score was incorporated into the model as a covariate, to calculate the propensity adjusted odds ratio (OR).

Likewise, a second propensity score was developed to adjust for the choice of initial steroid regimen.

All statistical analyses were performed using SPSS system (version 26.0 for Windows, SPSS Inc., Chicago, IL, USA). The statistical significance level was set at a two-sided p value of < 0.05. A hazard ratio (HR) or an odds ratio (OR) was reported along with 95% confidence interval (CI).

### 7. Ethics

The study was approved by the Institutional Review Board (CEIm) at Hospital Universitario Puerta de Hierro-Majadahonda (BRA-COR-2020–03), and a waiver for the informed consent was granted. The study complied with the provisions in EU and Spanish legislation on data protection and the Declaration of Helsinki 2013.

### 9. Registration

The protocol of the study was registered in EU PAS Register #EUPAS34753 on 15th April 2020 and publicly available at: http://www.encepp.eu/encepp/studySearch.htm

## RESULTS

During the study period, 848 patients with COVID-19 and pneumonia were admitted to the hospital. 463 out of 848 patients (55%) were included. Among them, 396 were treated with steroids, whilst 67 were assigned to the control cohort. A total of 385 patients were excluded from participation. Most of these patients were hospitalized with COVID-19 infection, but did not develop ARDS or increase in inflammatory markers.

### Clinical characteristics

Clinical characteristics of the cases and controls are displayed in Table 1. Median time to steroid treatment from the onset of symptoms was 10 days (IQR 8-13). Among patients treated with steroids, 310 (78.3 %) patients were initially treated with 1 mg/kg/day methylprednisolone or equivalent (22.5% of them received steroid pulses later-on) and 86 (21.7%) received pulses from the beginning.

**Table 1.** Baseline demographic and clinical characteristics of the patients in both cohorts.

| **N=463** | **STERIODS Cohort**<br>**(N=396)** | **CONTROL**<br>**Cohort**<br>**(N=67)** | **p value** |
| --- | --- | --- | --- |
| Gender (Men), n (%) | 276 (69.7) | 41 (61.2) | 0.200 |
| Age, Mean (SD) | 65.4 (12.9) | 68.1 (15.7) | 0.132 |
| Charlson score, Mean (SD) | 2.0 (2.3) | 2.3 (2.6) | 0.389 |
| Underlying Medical Conditions, n (%) | 306 (77.3) | 53 (79.1) | 0.874 |
| High Blood pressure | 182 (46.0) | 32 (47.8) | 0.793 |
| Ischemic heart disease | 72 (18.2) | 12 (17.9) | 0.957 |
| Diabetes | 84 (21.2) | 13 (19.4) | 0.871 |
| Obesity | 29 (7.3) | 6 (9.0) | 0.619 |
| Dyslipidemia | 113 (28.5) | 22 (32.8) | 0.471 |
| Cardiovascular risk factors | 249 (63.2) | 48 (71.6) | 0.215 |
| Chronic kidney disease | 24 (6.1) | 4 (6.0) | 0.977 |
| Onco-hematologic | 49 (12.4) | 16 (23.9) | **0.021** |
| COPD | 71 (17.9) | 10 (14.9) | 0.607 |
| Transplant (SOT/SCT) | 9 (2.3) | 1 (1.5) | 0.685 |
| Neurologic | 35 (8.8) | 11 (16.4) | 0.074 |
| Rheumatologic | 14 (3.5) | 1 (1.5) | 0.707 |
| Hepatic | 10 (2.5) | 5 (7.5) | 0.051 |
| Peptic Ulcer disease | 3 (0.8) | 3 (4.5) | **0.013** |
| Thromboembolic disease | 5 (1.3) | 3 (4.5) | 0.062 |
| Thyroid disorders | 15 (3.8) | 5 (7.5) | 0.189 |
| Immunosuppression | 37 (9.4) | 4 (6.0) | 0.171 |
| Clinical symptoms at admission, n (%) |  |  |  |
| Cough | 312 (79.6) | 44 (65.7) | **0.017** |
| Fever | 353 (90.3) | 56 (83.6) | 0.131 |
| Dyspnea | 272 (69.2) | 42 (62.7) | 0.321 |
| Gastrointestinal | 92 (23.2) | 13 (19.4) | 0.532 |
| Sore throat | 22 (5.6) | 4 (6.0) | 0.780 |
| Anosmia/ageusia | 28 (7.1) | 5 (7.5) | 0.802 |
| Myalgia | 82 (20.7) | 12 (17.9) | 0.743 |
| Headache | 29 (7.3) | 4 (6.0) | 0.691 |
| Fatigue | 63 (15.9) | 15 (22.4) | 0.216 |
| Chest pain | 23 (5.8) | 3 (4.5) | 0.662 |
| Rash | 2 (0.5) | 0 | 0.560 |
| Increased sputum production | 6 (1.5) | 5 (7.5) | **0.013** |
| Confusion | 18 (4.7) | 10 (15.4) | **0.003** |
| Days from onset of symptoms to diagnosis, Mean (SD) | 8.5 (5.1) | 6.9 (3.9) | **0.021** |
| Days from onset of symptoms to hospital admission, Mean (SD) | 7.6 (4.2) | 7.0 (3.7) | 0.231 |
| Days from onset of symptoms to therapy, Mean (SD) | 7.4 (4.1) | 7.1 (3.6) | 0.506 |
| Days from onset of symptoms to inclusion, Mean (SD) | 10.8 (4.8) | 8.7 (4.4) | **0.002** |
| Treatment, n (%) |  |  |  |
| Hydroxychloroquine | 393 (99.5) | 62 (92.5) | **0.001** |
| Lopinavir/Ritonavir | 287 (73.0) | 42 (62.7) | 0.106 |
| Azithromycin | 208 (53.9) | 29 (43.9) | 0.144 |
| Interferon | 186 (47.6) | 28 (41.8) | 0.427 |
| Tocilizumab | 177 (44.9) | 12 (18.5) | **< 0.001** |
| Anakinra | 8 (2.0) | 0 | 0.241 |
| Other treatments* | 65 (16.4) | 20 (29.9) | **0.009** |
| PaO<sub>2</sub>/FiO<sub>2</sub>, Mean (SD) | 263 (112.1) | 267 (78.9) | 0.878 |
| SatO<sub>2</sub>/FiO<sub>2</sub>, Mean (SD) | 286 (123.0) | 244 (91.9) | **0.021** |
| Brescia-COVID19 $\geq 2$, n (%) | 77 (17.4) | 16 (23.9) | 0.411 |
| ARDS, n (%) |  |  |  |
| No | 156 (39.4) | 9 (13.4) | **< 0.001** |
| Mild | 96 (24.2) | 43 (64.2) |  |
| Moderate | 116 (29.3) | 15 (22.4) |  |
| Severe | 28 (7.1) | 0 |  |
| Admitted to ICU at D0 | 30 (7.6%) | 0 | **0.013** |
| Laboratory (day 0), Mean (SD) |  |  |  |
| Lymphocyte counts | 1,004 (1,354) | 1,190 (1,042) | 0.342 |
| Lactate dehydrogenase | 396 (154) | 338 (117) | **0.018** |
| D- Dimer | 2.5 (7.6) | 2.1 (4.6) | 0.741 |
| C-reactive protein | 141 (85) | 122 (76) | 0.157 |
| Ferritin | 1,353 (2.220) | 763 (1.008) | 0.347 |
| Interleukin 6 (**IL-6**) | 196 (228) | 62 (62) | **0.039** |
| Chest CT (at hospital admission), n (%) |  |  |  |
| Normal imaging | 16 (4.1) | 2 (3.1) |  |
| Unilateral pneumonia | 29 (7.5) | 12 (18.5) |  |
| Bilateral interstitial pneumonia | 217 (55.8) | 27 (41.5) | **0.033** |
| Patchy bilateral pneumonia | 93 (23.9) | 16 (24.6) |  |
| Confluent bilateral pneumonia | 34 (8.7) | 8 (12.3) |  |

\*Including DRVr/ doxycycline/ clarithromycin and other antibiotics
COPD: chronic obstructive pulmonary disease; SOT: solid organ transplantation; SCT: stem cell transplantation; D0: day 0; SD: standard deviation; N/A: non-applicable; PaO<sub>2</sub>/FiO<sub>2</sub>: arterial oxygen tension/inspiratory oxygen fraction; SatO<sub>2</sub>/FiO<sub>2</sub>: oxygen saturation /inspiratory oxygen fraction; ARDS: acute respiratory distress syndrome; ICU: intensive care unit; Brescia-COVID19: respiratory severity scale Brescia-COVID-19; CT: computed tomography scan; IL-6: Interleukin 6; DRVr: ritonavir-boosted darunavir.

Patients treated with steroid pulses received a median of 3 pulses (IQR 2-4), followed by tapering in 25% of cases. Pulses of methylprednisolone were classified in the following groups: <250 mg/d (20.1%), 250 mg/d (62.5%), and 500 mg/d (17.1%).

### In-hospital mortality of patients treated with steroids compared to patients not treated with steroids

Global in-hospital mortality was 15.3%. Characteristics of survivors and non-survivors are shown in Appendix Table 1.

In-hospital mortality was lower in patients treated with steroids than in controls (13.9% versus 23.9%, OR 0.51 [0.27-0.96], p= 0.044) (Table 2). Steroid treatment reduced mortality by 41.8% relative to no steroid treatment (RRR 0.42 [0.048 to 0. 65]). We calculated a NNT (number necessary to treat) of 10. A propensity score to reduce the effect of steroid treatment selection bias was developed. Significant differences in baseline characteristics between steroid-treated and non-treated patients, such as onco-hematologic underlying conditions, peptic ulcer disease, LDH and SpO2, were considered for the propensity score. The difference in mortality persisted after applying the propensity score adjusted for steroid treatment (Table 2). Figure 1 demonstrates differences in the probability for survival at day 30 for patients with SARS-CoV2 infection, according to steroid treatment (log-rank p <0.001). Among patients with moderate or severe ARDS, in-hospital mortality was lower in patients treated with steroids than in the controls (26.2% versus 60%, OR 0.23 [0.08-0.71], p=0.014). Appendix Figures 1 and 2 show differences in probability of survival at day 30 according to steroid treatment, stratified according to ARDS severity.

**Figure 1.**
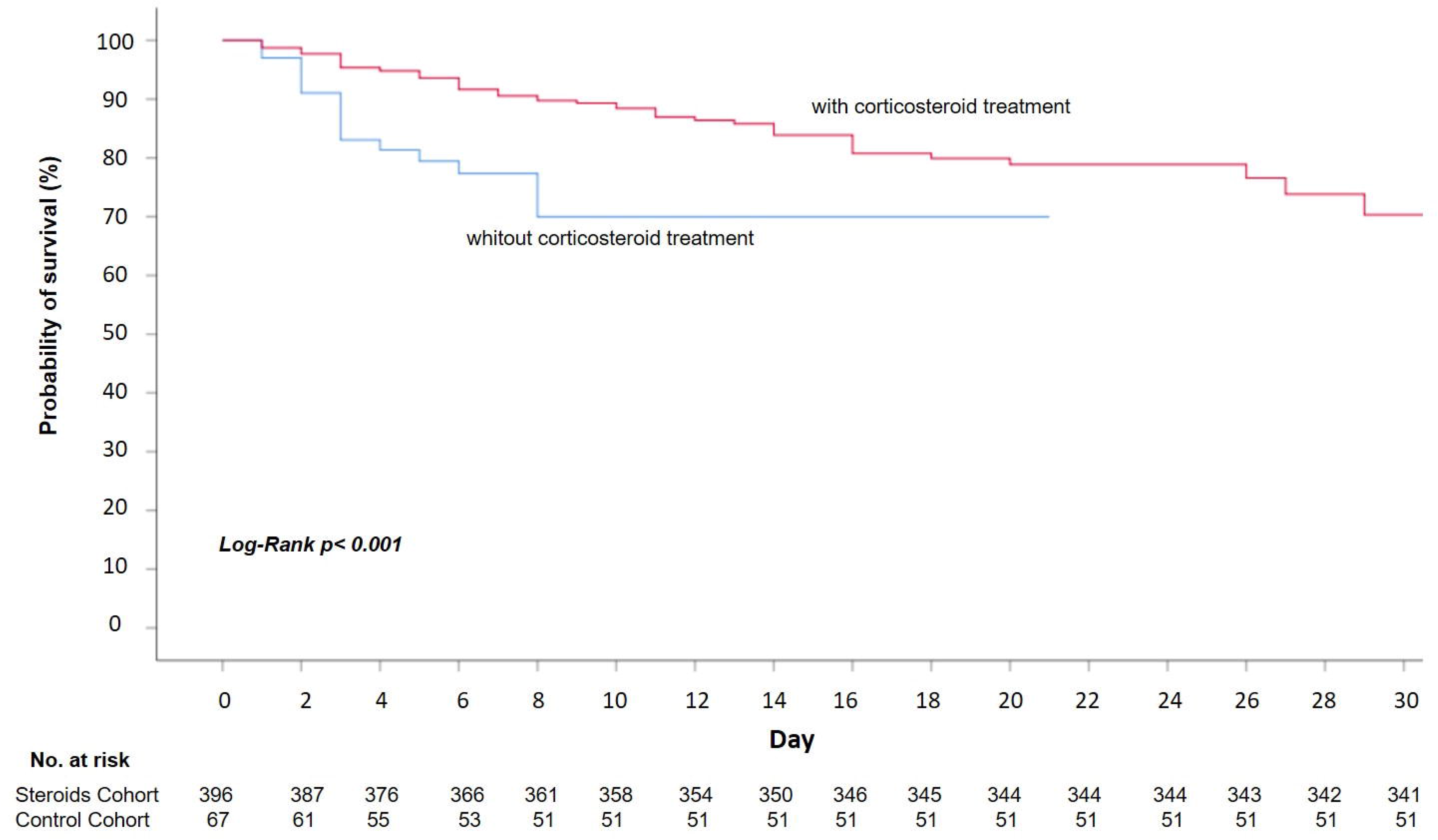
Probability of survival from D0 to hospital discharge of patients with SARS-COV-2 infection, according to steroid exposure.

The effect of steroid treatment on mortality in different subsets of patients was consistent with a protective effect, as can be seen in Figure 2.

**Figure 2.**
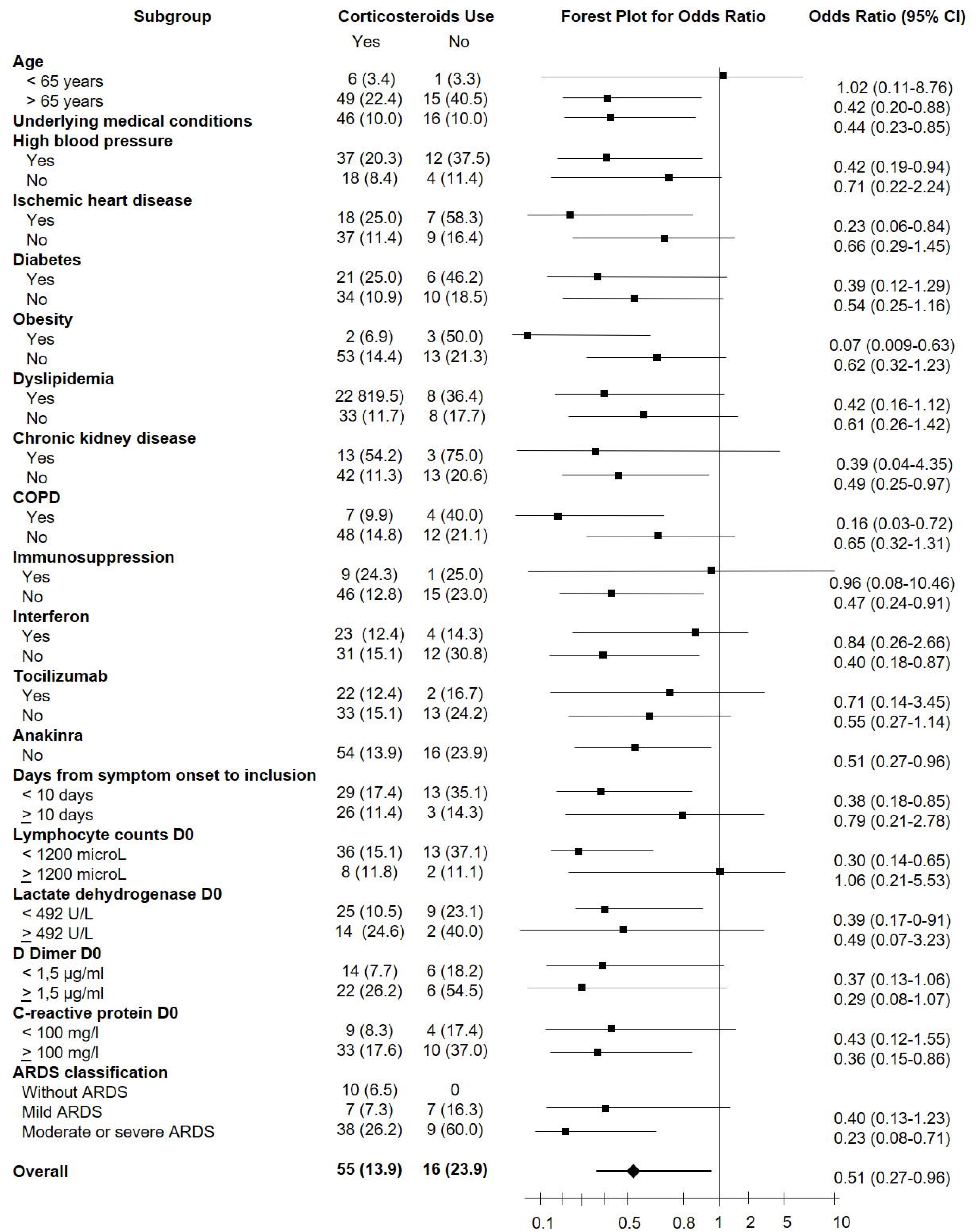
Forest plot of stratified analyses for in-hospital mortality showing the adjusted odds ratio of corticosteroid treatment. The subgroups were classified by demographic and disease characteristics.

**Table 2.** Association between steroid treatment and mortality in patients with SARS-COV-2 Infection, according to steroid exposure and steroid regimen.

|  | Survivors | Non-survivors |  |  |
| --- | --- | --- | --- | --- |
| Steroid Exposure | (N = 392) | (N = 71) | OR (95% CI) | p value |
| No corticosteroid treatment | 51 (76.1%) | 16 (23.9%) | 0.514 (0.274-0.965) | **0.038\*** |
| Steroid treatment | 341 (86.1%) | 55 (13.9%) |  |  |
| Steroid treatment (adjusted by PSM) |  |  | 0.360 (0.139-0.932) | **0.035\*** |
| **Steroid Regimen** |  |  |  |  |
| 1 mg/kg/day | 268 (86.5%) | 42 (13.5%) | 0.880 (0.449-1.726) | 0.710** |
| Pulses | 73 (84.9%) | 13 (15.1%) |  |  |

\*No steroids compared to steroid treatment (any regimen); \*\*Initial 1mg/kg/day compared to initial steroid pulses
PSM: propensity score matching

Table 3 shows the risk factors for mortality in both univariable and multivariable analyses, including those adjusted by the propensity score for steroid treatment. Older age, chronic kidney disease, more severe ARDS and elevated lactate dehydrogenase (LDH) levels were independent risk factors for mortality, whereas steroid treatment was an independent protective factor. Except for ARDS severity, these results were confirmed when adjusted by propensity score for steroid treatment.

**Table 3.**
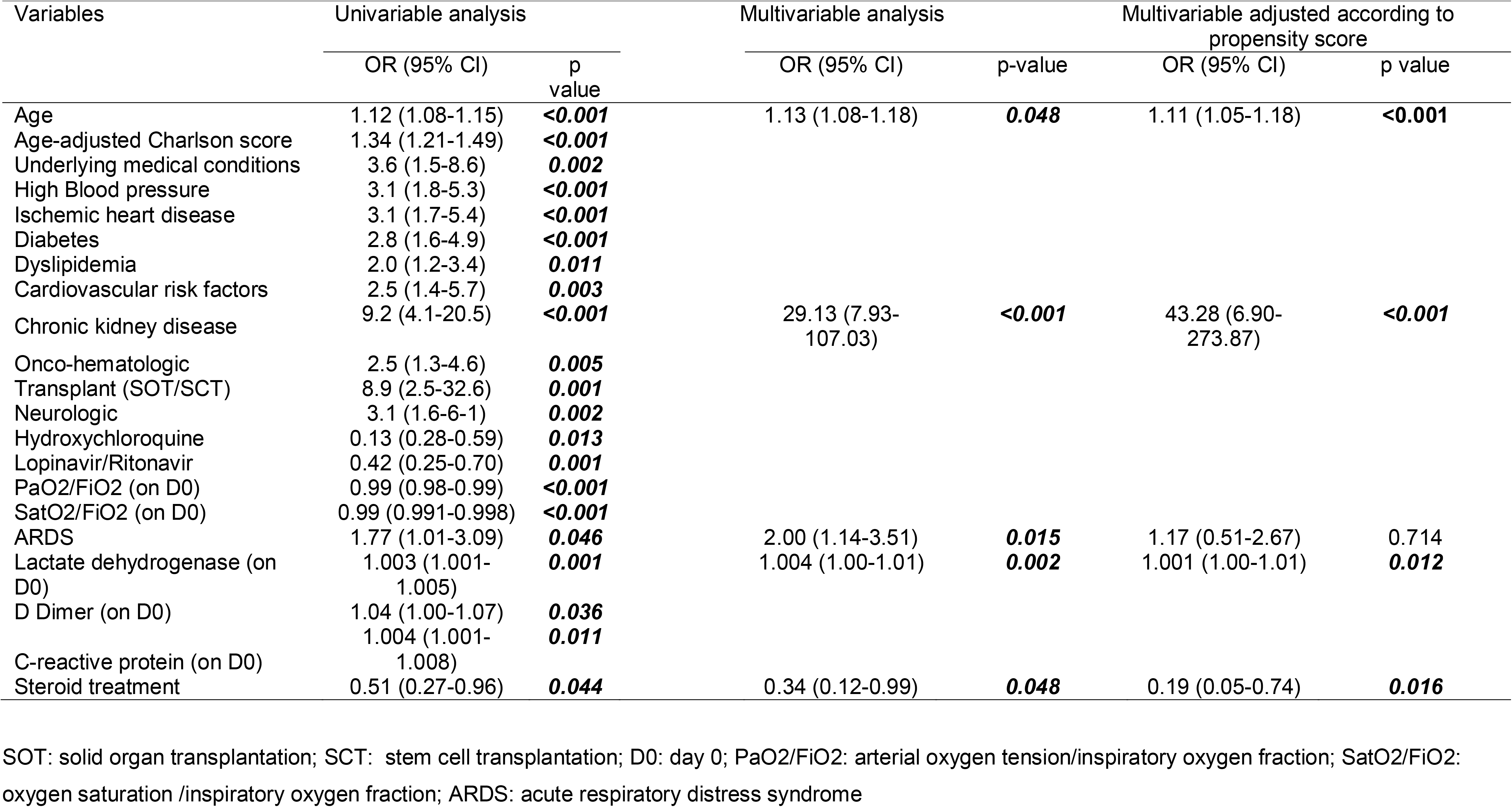
Univariable and multivariable analyses of factors associated with hospital mortality in patients with SARS-COV-2 Infection.

### In-hospital mortality of patients treated with different steroid regimens: pulses versus 1 mg/kg/day

Characteristics of patients initially treated with 1 mg/kg/day of methylprednisolone (or equivalent) versus those initially treated with steroid pulses are displayed in Appendix Table 2. Being treated with either regimen was not associated with in-hospital mortality (13.5% versus 15.1%, OR 0.880 [0.449-1.726], p=0.710; RRR 0.10 [-0.59 to 0.50]).

A propensity score for the choice of initial steroid regimen was developed. After adjusting by this propensity score, there were still no differences in mortality. <u>Characteristics of patients initially treated with 1 mg/kg/day that eventually required salvage steroid pulses</u>

A subset of patients initially treated with 1 mg/kg/d of methylprednisolone received subsequent steroid pulses, in a median of 3-days-time (IQR 2-7). Baseline characteristics of these patients, as opposed to those who did not, are presented in Appendix Table 3. Diabetic patients, those with underlying neurologic disease or higher levels of LDH at steroid initiation were more prone to require subsequent pulses, according to the multivariable analysis.

## DISCUSSION

Our results show that survival of patients with SARS-CoV2 pneumonia is higher in patients treated with steroids than in those not treated. These results support the use of steroids in SARS-CoV2 infection. In-hospital mortality was not different between initial regimens of 1 mg/kg/day of methylprednisolone and steroid pulses.

The timing of the steroid administration might be decisive. In the present study, patients received steroid treatment in a median of 10 days after onset of symptoms, presumably during the inflammatory phase of the disease. Three distinct stages of COVID-19 illness have been suggested^1^. Siddiqi et al^1^ suggested that from stage IIB-on, starting when hypoxia develops, anti-inflammatory therapies, such as steroids, could be beneficial, due to the predominant role of inflammation in its pathophysiology.

The warning against the use of steroids in COVID19 is based on studies that administered this therapy earlier-on during the course of the disease, and relies on the experience from different viruses^12^. Moreover, it has been speculated that steroid administration in patients with SARS-CoV2 infection could be deleterious due to an increase of viral shedding or a delay in viral clearance. Although this theory was not confirmed in a recent work by Fang^13^, it is worth considering if steroids are to be administered early-on in the course of the disease.

In the present series, steroids were used in patients with hypoxemia that were not at an early phase of the disease, as stated by the median time from the onset of symptoms to steroid administration. As many as 64% of the cases fulfilled ARDS criteria at the time of steroid administration. At this stage, as suggested by the lower rates of mortality seen in the treatment cohort when compared to the control group, steroid treatment was beneficial. In general, guidelines recommend not using corticosteroids in patients with COVID-19, or using them only in intubated patients^14^ or in the setting of randomized clinical trials^15^. In our series, steroid treatment was beneficial in patients with moderate to severe ARDS, but a trend to a better survival was also seen in cases with mild ARDS, though it did not reach statistical significance, possibly due to a small sample size. When steroids are delayed to more advanced stages, we might be missing a therapeutic window to prevent the evolution to severe ARDS and the need for mechanical ventilation. Nevertheless, the optimal stage for steroid treatment remains to be elucidated.

Patients with higher LDH levels responded better to steroid treatment in the present series. As LDH can be considered a surrogate marker for the extent of lung involvement, these results would indicate that patients with more extensive lung damage might benefit more from steroid treatment. In this respect, our results are in line with those reported by others^5^.

A pattern of cytokines resembling that of secondary hemophagocytic lymphohistiocytosis has been associated with SARS-CoV2 infection^16^. Mehta et al^17^ suggested a role for corticosteroid in patients with severe COVID-19 and hyperinflammation diagnosed based on cytokine elevation profile. In our series, steroid protective effect was more intense in cases with inflammatory marker abnormalities such as higher D-dimer and C-reactive protein levels.

Optimal steroid dosing also needs clarification. Most patients treated with steroids in the present series received a weight-adjusted dose, but a significant proportion of patients (39.4%) received higher doses in pulses, either from the start or as salvage therapy, after a weight-adjusted course. In our series, we were not able to demonstrate a difference in mortality between these two regimens, even after adjusting by a propensity score taking into consideration the regimen choice and disease severity. An analysis of secondary adverse effects, which are usually dose-related, would help decide between regimens, if both dosing regimens were confirmed to be associated with equivalent outcomes.

Aiming to clarify which patients would eventually end-up requiring salvage steroid pulse therapy, we compared baseline characteristics of those starting with 1 mg/kg/day regimen. Our results suggest that diabetic patients and those with underlying neurologic conditions or higher levels of LDH could benefit from an earlier administration of steroid pulses, considering that they seem to be more prone to require them later-on. These results should be confirmed in further studies.

Our results are in line with a more preliminary work by Wang^18^, who reported a shorter duration of fever and a faster improvement of SpO2 in cases of severe SARS-CoV2 pneumonia treated with 1-2 mg/kg/day of methylprednisolone during a period of 5-7 days. The present study has a considerably larger sample size, a more diverse population, and the added value of a propensity score to adjust for steroid treatment. Moreover, we report an impact on in-hospital mortality.

A study by Zhou^8^, including only critical patients and without a control group, suggested that steroid treatment could enhance oxygen saturation and arterial PaO2/ FiO2 ratio, although mortality remained similar to that reported in the literature. Our study suggests that besides ICU patients with severe ARDS, other subsets of patients in an earlier phase of the disease could benefit from steroid therapy, and possibly avoid ICU admission.

The present study is a retrospective study that analyses real-life data, and as such, treated and untreated patients are not comparable according to all baseline characteristics. To overcome this limitation, we applied two propensity scores to the analysis, one for steroid treatment versus no steroid treatment and the second one, for the initial steroid regimen choice. Results were confirmed when including the propensity scores. As a single center study, the results need external validation. The only outcome that was evaluated in the study was mortality. We consider that ICU admission during the study period is not a reliable marker of poor outcome, given the scarcity of available ICU beds during that critical moments of the pandemic, which forced to apply strict restrictions for ICU admission.

The potential impact of steroids in the mortality of COVID-19 pneumonia suggested by this study supports the need to carry out randomized clinical trials with the aim to establish their role. The optimal timing for administration, the subset of patients with the best risk/benefit ratio and the appropriate dosing and duration remain to be elucidated.

## Data Availability

After publication, the data will be made available to others on reasonable requests to the corresponding authors. A proposal with a detailed description of study objectives and statistical analysis plan will be needed for evaluation of the reasonability of requests. Additional materials might also be required during the process of evaluation. De-identified participant data will be provided after approval from the principal researchers of the Hospital Puerta de Hierro-Majadahonda.

## Authors contribution

Conceptualization and study design: AFC, BRA

Methodology: BRA, AFC, ASL, AMG; CAS

Data collection: AMG, ASL, GCS, PMS, LGG, SBA, AGG, AVA, JGI, IMT, ESC, CPH, LDTC

Data interpretation: BRA, AFC, EMR, ARM, ACD

Writing first draft: AFC, BRA

Critical revision for important intellectual content, all authors; final approval, all authors;

Revision and English proofreading: CPH

All authors agree to be accountable for all aspects of the work in ensuring that questions related to the accuracy or integrity of any part of the work are appropriately investigated and resolved.

AFC and BRA had full access to all the data in this study and take complete responsibility for the integrity of the data and the accuracy of the data analysis.

## Other COVID19-Steroids Study Group Collaborators

see appendix

## Disclaimer

The results, discussion and conclusions are from the authors and do not necessarily represent the position of the Institution.

## ABBREVIATIONS

SARS-CoV: severe acute respiratory syndrome coronavirus
COVID-19: coronavirus disease 2019
Steroids: glucocorticoids or corticoids
OR: Odds ratio
MERS-CoV: Middle east respiratory syndrome coronavirus
PaO2/FiO2: arterial oxygen tension/inspiratory oxygen fraction
CT: computed tomography
ARDS: acute respiratory distress syndrome
CRP: C-reactive protein
PEEP: positive end expiratory pressure
SD: standard deviation
CI: confidence interval
PSM: propensity-score matching
RRR: relative risk reduction
NNT: number necessary to treat
HR: hazard ratio
IQR: interquartile range
COPD: chronic obstructive pulmonary disease
LDH: Lactate dehydrogenase
SpO2: plasma oxygen saturation
ICU: Intensive care Unit

---

**Appendix Table 1.**
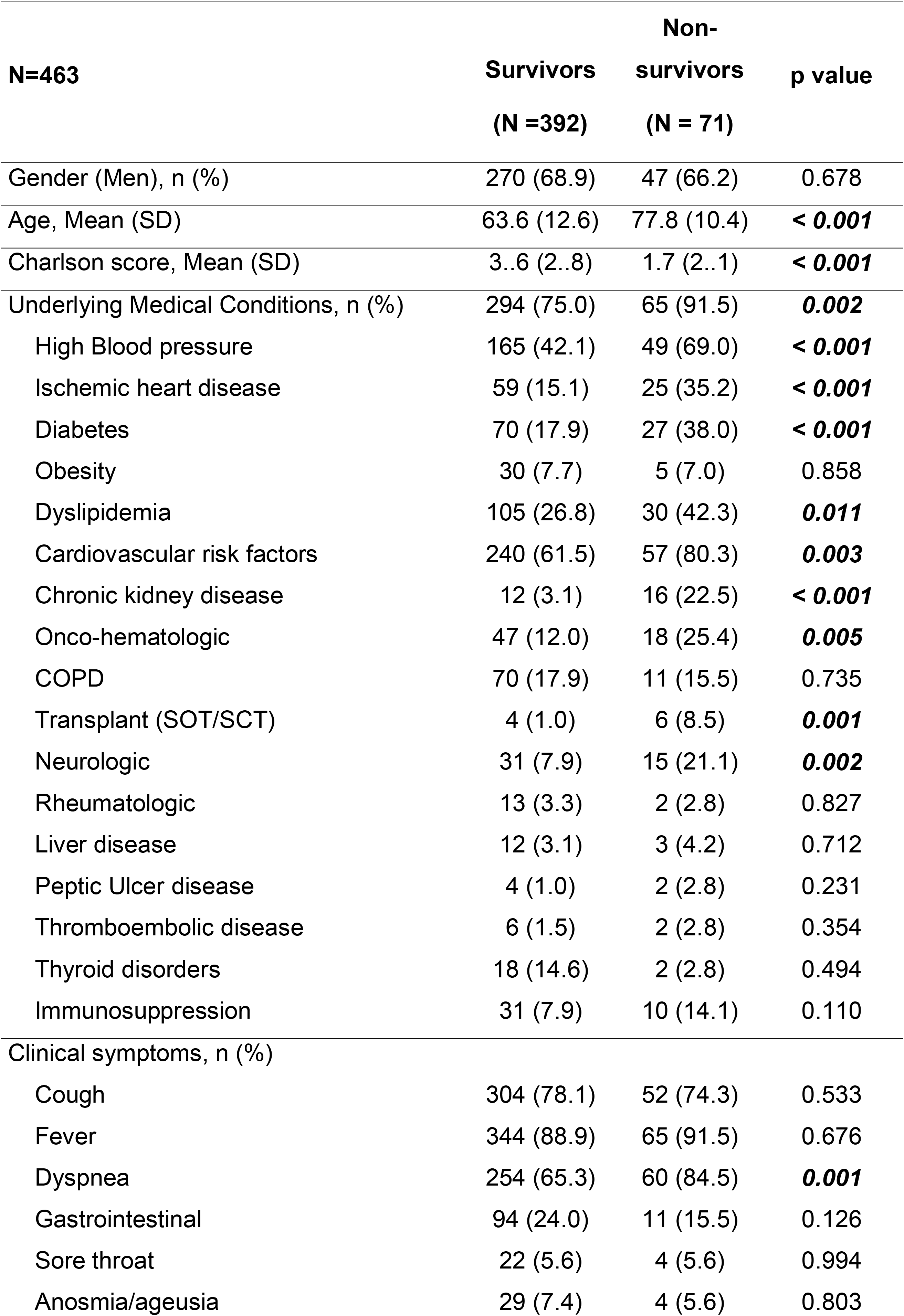
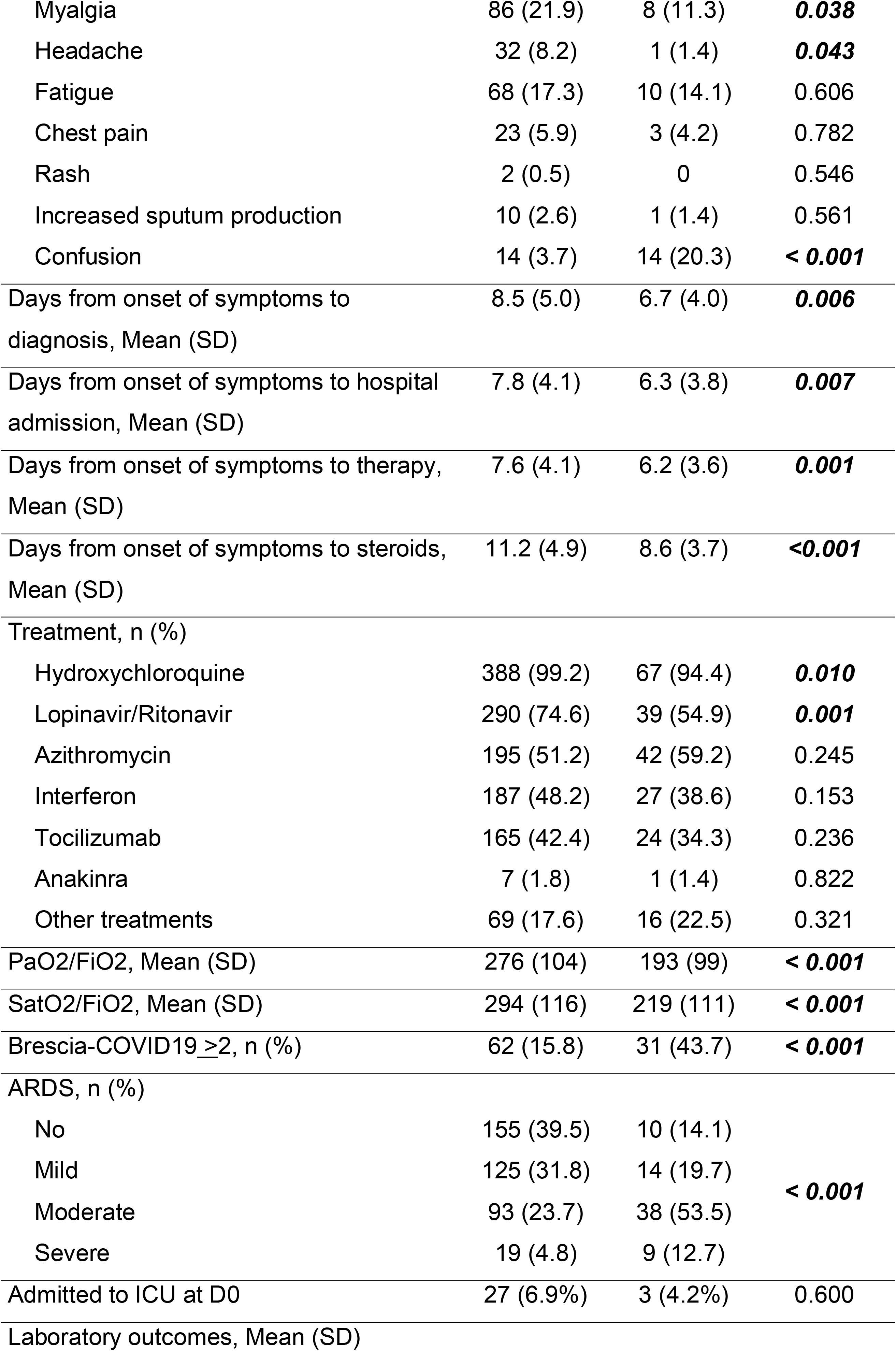
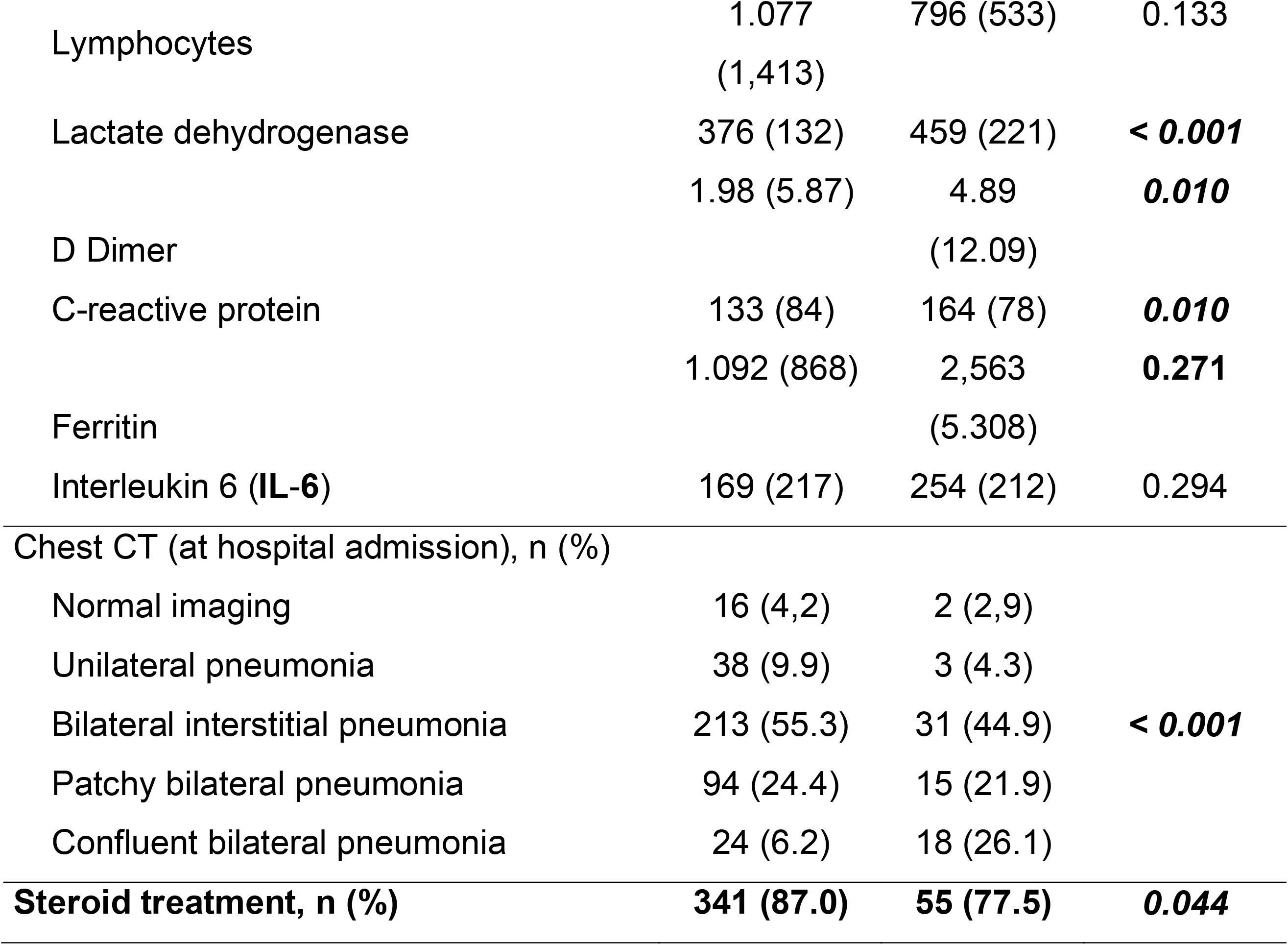
Comparison between survivors and non-survivors

**Appendix Table 2.** Baseline demographic and clinical characteristics of the patients according to initial steroid regimen

| **N=396** | **1 mg/kg/day Cohort**<br>**(N=310)** | **Pulses Cohort**<br>**(N=86)** | **p value** |
| --- | --- | --- | --- |
| Gender (Men), n (%) | 213 (68.7%) | 63 (73.3%) | 0.417 |
| Age, Mean (SD) | 65.3 (13.0) | 65.8 (12.2) | 0.775 |
| Charlson score, Mean (SD) | 2.03 (2.2) | 2.00 (2.3) | 0.916 |
| Underlying Medical Conditions, n (%) |  |  |  |
| High Blood pressure | 148 (47.7%) | 34 (39.5%) | 0.177 |
| Ischemic heart disease | 63 (20.3%) | 9 (10.5%) | **0.036** |
| Diabetes | 65 (21%) | 19 (22.1%) | 0.821 |
| Obesity | 24 (7.7%) | 5 (5.8%) | 0.544 |
| Dyslipidemia | 90 (29%) | 23 (26.7%) | 0.678 |
| Cardiovascular risk factors | 204 (66.2%) | 45 (52.3%) | **0.018** |
| Chronic kidney disease | 19 (6.1%) | 5 (5.8%) | 0.914 |
| Onco-hematologic | 34 (11%) | 15 (17.4%) | 0.107 |
| COPD | 53 (17.1%) | 18 (20.9%) | 0.412 |
| Transplant (SOT/SCT) | 8 (2.6%) | 1 (1.2%) | 0.435 |
| Neurologic | 24 (7.7%) | 11 (12.8%) | 0.144 |
| Rheumatologic | 8 (2.6%) | 6 (7%) | 0.051 |
| Liver disease | 8 (2.6%) | 2 (2.3%) | 0.894 |
| Peptic Ulcer disease | 2 (0.6%) | 1 (1.2%) | 0.624 |
| Thromboembolic disease | 4 (1.3%) | 1 (1.2%) | 0.925 |
| Thyroid disorders | 11 (3.5%) | 4 (4.7%) | 0.636 |
| Immunosuppression | 29 (9.4%) | 8 (9.3%) | 0.981 |
| Clinical symptoms at admission, n (%) |  |  |  |
| Cough | 245 (80.1%) | 67 (77.9%) | 0.661 |
| Fever | 281 (91.5%) | 72 (85.7%) | 0.111 |
| Dyspnea | 219 (71.3%) | 53 (61.6%) | 0.085 |
| Gastrointestinal | 75 (24.2%) | 17 (19.8%) | 0.390 |
| Sore throat | 18 (5.8%) | 4 (4.7%) | 0.679 |
| Anosmia/ageusia | 20 (6.5%) | 8 (9.3%) | 0.362 |
| Myalgia | 66 (21.3%) | 16 (18.6%) | 0.587 |
| Headache | 25 (8.1%) | 4 (4.7%) | 0.282 |
| Fatigue | 44 (14.2%) | 19 (22.1%) | 0.076 |
| Chest pain | 17 (5.5%) | 6 (7%) | 0.600 |
| Rash | 2 (0.6%) | 0 (0%) | 0.455 |
| Increased sputum production | 3 (1%) | 3 (3.5%) | 0.090 |
| Confusion | 10 (3.3%) | 8 (9.4%) | **0.018** |
| Days from onset of symptoms to diagnosis, Mean (SD) | 8.5 (5.4) | 8.5 (4.0) | 0.985 |
| Days from onset of symptoms to hospital admission, Mean (SD) | 7.5 (4.2) | 8.3 (4.1) | 0.118 |
| Days from onset of symptoms to therapy, Mean (SD) | 7.4 (4.2) | 7.7 (3.8) | 0.599 |
| Days from onset of symptoms to steroids, mean (SD) | 10.7 (5.0) | 11.32 (4.0) | 0.307 |
| Number of Pulses, Mean (SD) | *3.3 (2.0) | 3.5 (2.0) | 0.522 |
| Pulse dose | * |  | **0.015** |
| <250 mg | 7 (10.3%) | 24 (15.8%) |  |
| 250 mg | 46 (67.6%) | 49 (51.6%) |  |
| > 500 mg | 15 (22.1%) | 11 (13.1%) |  |
| Days from steroid 1mg/kg/d to pulses*, Mean (SD) | 4.9 (3.7) | N/A |  |
| Treatment, n (%) |  |  |  |
| Hydroxychloroquine | 307 (99.4%) | 86 (100%) | 0.454 |
| Lopinavir/Ritonavir | 222 (72.1%) | 65 (76.5%) | 0.419 |
| Azithromycin | 159 (52.8%) | 49 (57.6%) | 0.431 |
| Interferon | 146 (47.9%) | 40 (46.5%) | 0.903 |
| -before or together with steroids | 138 (93.9%) | 33 (94.3%) | 0.999 |
| -after steroids | 9 (6.1%) | 2 (5.7%) |  |
| Tocilizumab | 130 (42.2%) | 47 (54.7%) | 0.05 |
| -before or together with steroids | 77 (59.7%) | 25 (59.5%) | 0.999 |
| -after steroids | 52 (40.3%) | 17 (40.5%) |  |
| Anakinra | 4 (1.3%) | 4 (4.7%) | 0.071 |
| -before or together with steroids | 2 (66.7%) | 0 | 0.4 |
| -after steroids | 1 (33.3%) | 3 (75%) |  |
| Other treatments** |  |  |  |
| PaO<sub>2</sub>/FiO<sub>2</sub>, Mean (SD) | 261.4 (110.4) | 270.5 (118.4) | 0.544 |
| SatO<sub>2</sub>/FiO<sub>2</sub>, Mean (SD) | 286.7 (123.6) | 300.6 (125.6) | 0.543 |
| Brescia-COVID19 $\geq 2$, n (%) | 62 (20%) | 15 (17.4%) | 0.596 |
| ARDS, n (%) |  |  | 0.110 |
| No | 115 (37.1%) | 40 (46.5%) |  |
| Mild | 82 (26.5%) | 14 (16.3%) |  |
| Moderate/ Severe | 113 (36.5%) | 32 (37.2%) |  |
| Admitted to ICU at D0 | 28 (9%) | 2 (2.3%) | 0.038 |
| Laboratory (day 0), Mean (SD) |  |  |  |
| Lymphocyte counts | 1040.6 (1522 (1) | 879.2 (412.0) | 0.384 |
| Lactate dehydrogenase | 396.7 (157.7) | 395.6 (141.5) | 0.963 |
| D- Dimer | 2.1 (6.4) | 3.6 (10.6) | 0.295 |
| C-reactive protein | 139.6 (84.0) | 145.7 (88.4) | 0.611 |
| Ferritin | 1486.8 (2589.2) | 1042.7 (783.6) | 0.340 |
| Interleukin 6 (IL-6) | 195.1 (230.3) | 201.1 (229.7) | 0.921 |
| Chest CT (at hospital admission), n (%) |  |  | 0.275 |
| Normal imaging | 14 (4.6%) | 2 (2.4%) |  |
| Unilateral pneumonia | 24 (7.9%) | 5 (5.9%) |  |
| Bilateral interstitial pneumonia | 169 (55.6%) | 48 (56.5%) |  |
| Patchy bilateral pneumonia | 75 (24.7%) | 18 (21.2%) |  |
| Confluent bilateral pneumonia | 22 (7.2%) | 12 (14.1%) |  |

\*In 70 (22.6%) patients that received steroid pulses after a 1 mg/kg/day regimen \*\*Including DRVr/ doxycycline/ clarithromycin and other antibiotics
COPD: chronic obstructive pulmonary disease; SOT: solid organ transplantation; SCT: stem cell
transplantation; D0: day 0; SD: standard deviation; N/A: non-applicable; PaO<sub>2</sub>/FiO<sub>2</sub>: arterial oxygen tension/inspiratory oxygen fraction; SatO<sub>2</sub>/FiO<sub>2</sub>: oxygen saturation /inspiratory oxygen fraction; ICU: intensive care unit; ARDS: acute respiratory distress syndrome; Brescia-COVID19: respiratory severity scale Brescia-COVID-19; CT: computed tomography scan; IL-6: Interleukin 6; DRVr: ritonavir-boosted darunavir

**Appendix Table 3.**
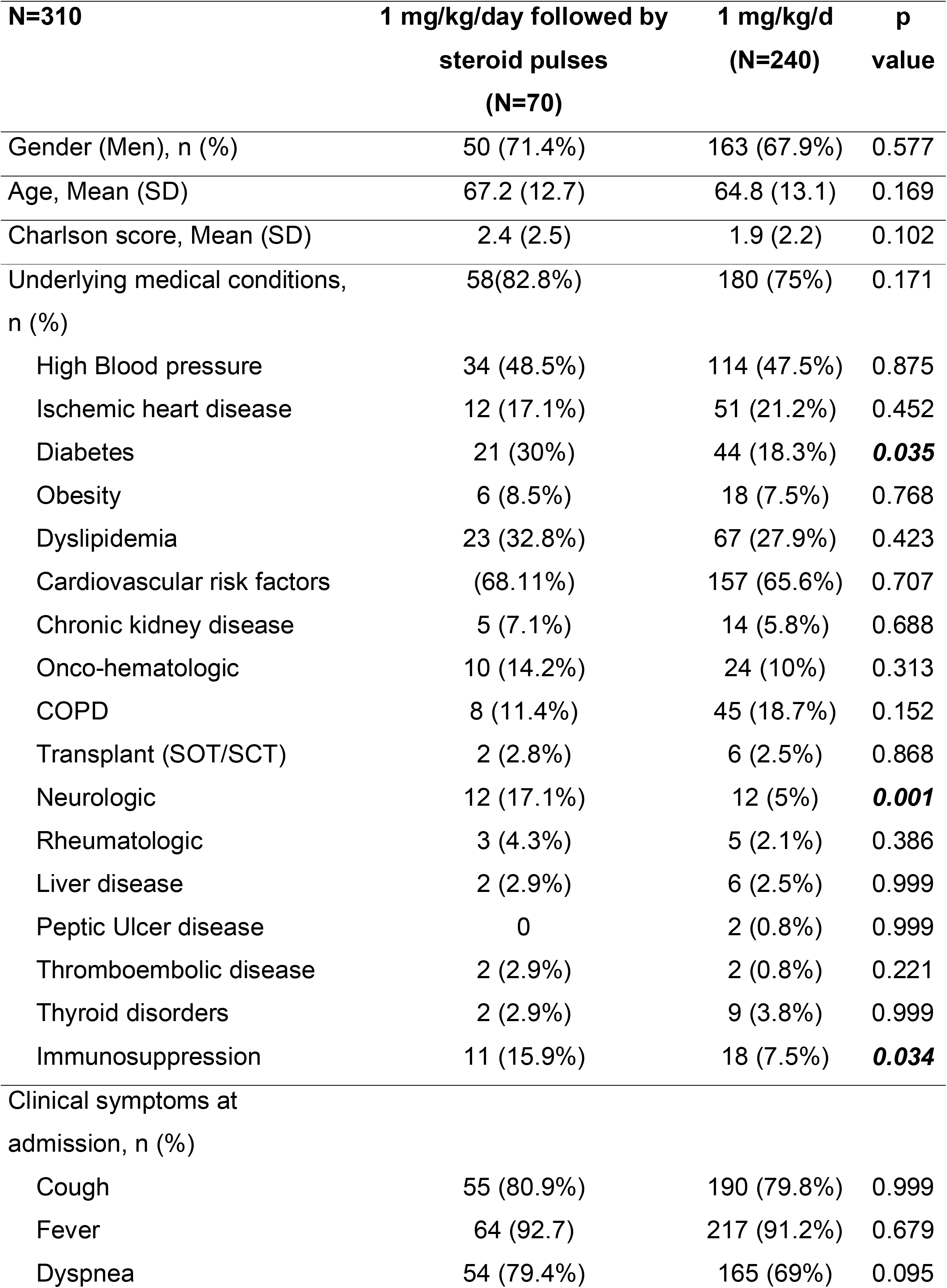
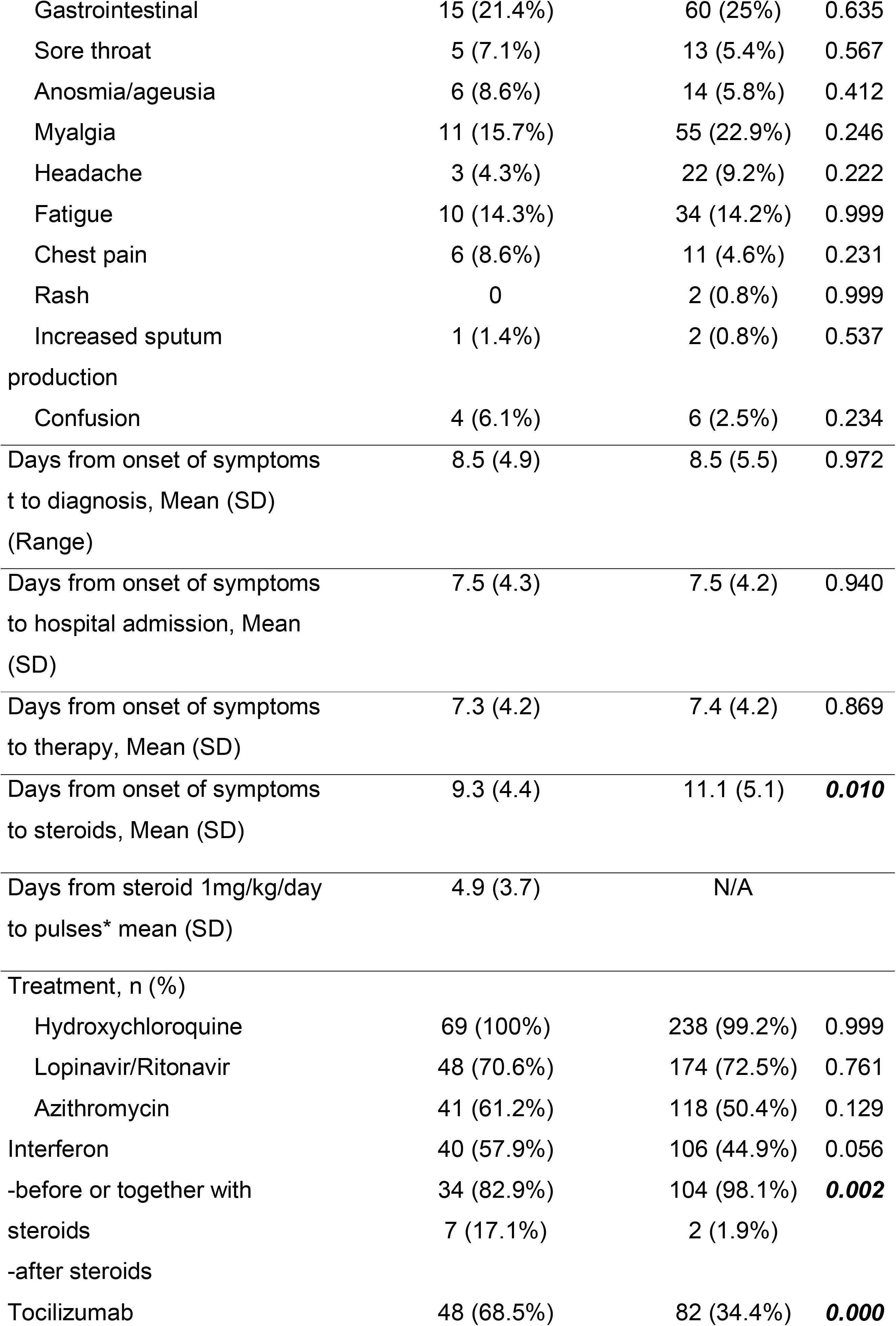
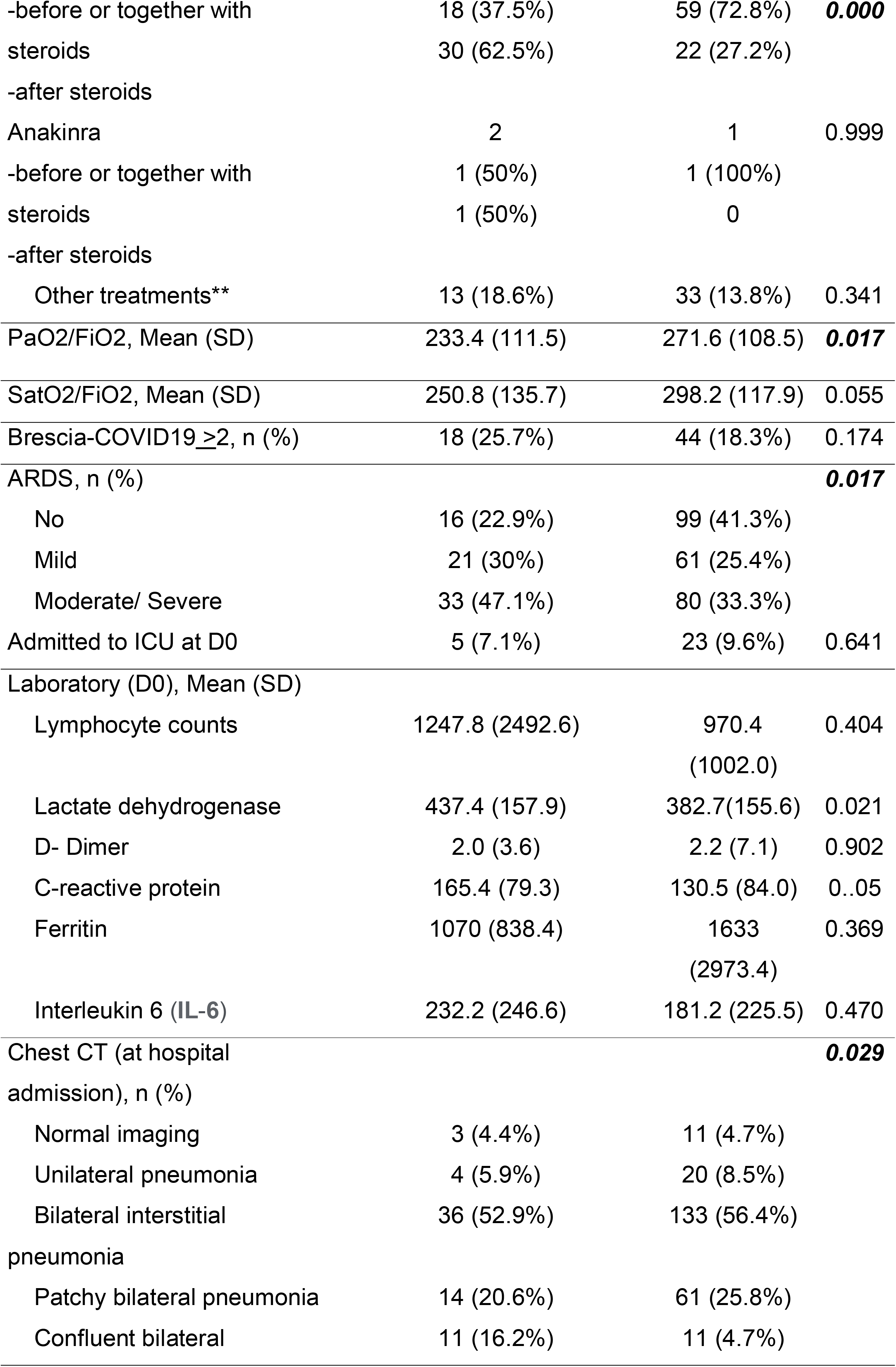
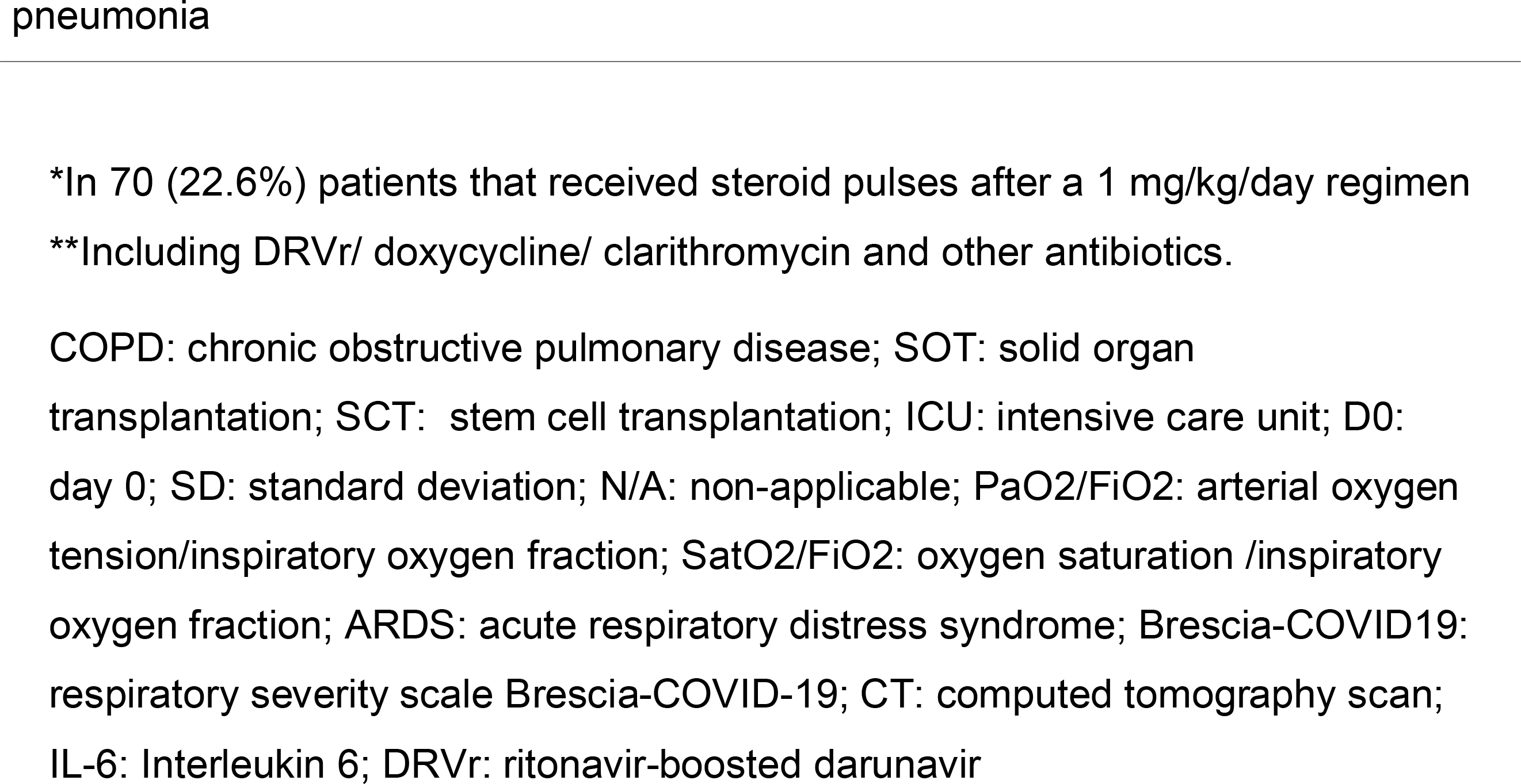
Baseline demographic and clinical characteristics of the patients initially receiving 1 mg/kg/day according to subsequent need for steroid pulses

| **N=310** | **1 mg/kg/day followed by**<br>**steroid pulses**<br>**(N=70)** | **1 mg/kg/d**<br>**(N=240)** | **p**<br>**value** |
| --- | --- | --- | --- |
| Gender (Men), n (%) | 50 (71.4%) | 163 (67.9%) | 0.577 |
| Age, Mean (SD) | 67.2 (12.7) | 64.8 (13.1) | 0.169 |
| Charlson score, Mean (SD) | 2.4 (2.5) | 1.9 (2.2) | 0.102 |
| Underlying medical conditions,<br>n (%) | 58(82.8%) | 180 (75%) | 0.171 |
| High Blood pressure | 34 (48.5%) | 114 (47.5%) | 0.875 |
| Ischemic heart disease | 12 (17.1%) | 51 (21.2%) | 0.452 |
| Diabetes | 21 (30%) | 44 (18.3%) | **0.035** |
| Obesity | 6 (8.5%) | 18 (7.5%) | 0.768 |
| Dyslipidemia | 23 (32.8%) | 67 (27.9%) | 0.423 |
| Cardiovascular risk factors | (68.11%) | 157 (65.6%) | 0.707 |
| Chronic kidney disease | 5 (7.1%) | 14 (5.8%) | 0.688 |
| Onco-hematologic | 10 (14.2%) | 24 (10%) | 0.313 |
| COPD | 8 (11.4%) | 45 (18.7%) | 0.152 |
| Transplant (SOT/SCT) | 2 (2.8%) | 6 (2.5%) | 0.868 |
| Neurologic | 12 (17.1%) | 12 (5%) | **0.001** |
| Rheumatologic | 3 (4.3%) | 5 (2.1%) | 0.386 |
| Liver disease | 2 (2.9%) | 6 (2.5%) | 0.999 |
| Peptic Ulcer disease | 0 | 2 (0.8%) | 0.999 |
| Thromboembolic disease | 2 (2.9%) | 2 (0.8%) | 0.221 |
| Thyroid disorders | 2 (2.9%) | 9 (3.8%) | 0.999 |
| Immunosuppression | 11 (15.9%) | 18 (7.5%) | **0.034** |
| Clinical symptoms at<br>admission, n (%) |  |  |  |
| Cough | 55 (80.9%) | 190 (79.8%) | 0.999 |
| Fever | 64 (92.7) | 217 (91.2%) | 0.679 |
| Dyspnea | 54 (79.4%) | 165 (69%) | 0.095 |
| Gastrointestinal | 15 (21.4%) | 60 (25%) | 0.635 |
| Sore throat | 5 (7.1%) | 13 (5.4%) | 0.567 |
| Anosmia/ageusia | 6 (8.6%) | 14 (5.8%) | 0.412 |
| Myalgia | 11 (15.7%) | 55 (22.9%) | 0.246 |
| Headache | 3 (4.3%) | 22 (9.2%) | 0.222 |
| Fatigue | 10 (14.3%) | 34 (14.2%) | 0.999 |
| Chest pain | 6 (8.6%) | 11 (4.6%) | 0.231 |
| Rash | 0 | 2 (0.8%) | 0.999 |
| Increased sputum<br>production | 1 (1.4%) | 2 (0.8%) | 0.537 |
| Confusion | 4 (6.1%) | 6 (2.5%) | 0.234 |
| Days from onset of symptoms<br>t to diagnosis, Mean (SD)<br>(Range) | 8.5 (4.9) | 8.5 (5.5) | 0.972 |
| Days from onset of symptoms<br>to hospital admission, Mean<br>(SD) | 7.5 (4.3) | 7.5 (4.2) | 0.940 |
| Days from onset of symptoms<br>to therapy, Mean (SD) | 7.3 (4.2) | 7.4 (4.2) | 0.869 |
| Days from onset of symptoms<br>to steroids, Mean (SD) | 9.3 (4.4) | 11.1 (5.1) | **0.010** |
| Days from steroid 1mg/kg/day<br>to pulses* mean (SD) | 4.9 (3.7) | N/A |  |
| Treatment, n (%) |  |  |  |
| Hydroxychloroquine | 69 (100%) | 238 (99.2%) | 0.999 |
| Lopinavir/Ritonavir | 48 (70.6%) | 174 (72.5%) | 0.761 |
| Azithromycin | 41 (61.2%) | 118 (50.4%) | 0.129 |
| Interferon | 40 (57.9%) | 106 (44.9%) | 0.056 |
| -before or together with<br>steroids | 34 (82.9%)<br>7 (17.1%) | 104 (98.1%)<br>2 (1.9%) | **0.002** |
| -after steroids |  |  |  |
| Tocilizumab | 48 (68.5%) | 82 (34.4%) | **0.000** |
| -before or together with steroids | 18 (37.5%)<br>30 (62.5%) | 59 (72.8%)<br>22 (27.2%) | **0.000** |
| -after steroids |  |  |  |
| Anakinra | 2 | 1 | 0.999 |
| -before or together with steroids | 1 (50%)<br>1 (50%) | 1 (100%)<br>0 |  |
| -after steroids |  |  |  |
| Other treatments** | 13 (18.6%) | 33 (13.8%) | 0.341 |
| PaO2/FiO2, Mean (SD) | 233.4 (111.5) | 271.6 (108.5) | **0.017** |
| SatO2/FiO2, Mean (SD) | 250.8 (135.7) | 298.2 (117.9) | 0.055 |
| Brescia-COVID19<sub>≥2</sub>, n (%) | 18 (25.7%) | 44 (18.3%) | 0.174 |
| ARDS, n (%) |  |  | **0.017** |
| No | 16 (22.9%) | 99 (41.3%) |  |
| Mild | 21 (30%) | 61 (25.4%) |  |
| Moderate/ Severe | 33 (47.1%) | 80 (33.3%) |  |
| Admitted to ICU at D0 | 5 (7.1%) | 23 (9.6%) | 0.641 |
| Laboratory (D0), Mean (SD) |  |  |  |
| Lymphocyte counts | 1247.8 (2492.6) | 970.4 (1002.0) | 0.404 |
| Lactate dehydrogenase | 437.4 (157.9) | 382.7(155.6) | 0.021 |
| D- Dimer | 2.0 (3.6) | 2.2 (7.1) | 0.902 |
| C-reactive protein | 165.4 (79.3) | 130.5 (84.0) | 0.05 |
| Ferritin | 1070 (838.4) | 1633 (2973.4) | 0.369 |
| Interleukin 6 (IL-6) | 232.2 (246.6) | 181.2 (225.5) | 0.470 |
| Chest CT (at hospital admission), n (%) |  |  | **0.029** |
| Normal imaging | 3 (4.4%) | 11 (4.7%) |  |
| Unilateral pneumonia | 4 (5.9%) | 20 (8.5%) |  |
| Bilateral interstitial pneumonia | 36 (52.9%) | 133 (56.4%) |  |
| Patchy bilateral pneumonia | 14 (20.6%) | 61 (25.8%) |  |
| Confluent bilateral | 11 (16.2%) | 11 (4.7%) |  |

\*In 70 (22.6%) patients that received steroid pulses after a 1 mg/kg/day regimen
\*\*Including DRVr/ doxycycline/ clarithromycin and other antibiotics.
COPD: chronic obstructive pulmonary disease; SOT: solid organ transplantation; SCT: stem cell transplantation; ICU: intensive care unit; D0: day 0; SD: standard deviation; N/A: non-applicable; PaO<sub>2</sub>/FiO<sub>2</sub>: arterial oxygen tension/inspiratory oxygen fraction; SatO<sub>2</sub>/FiO<sub>2</sub>: oxygen saturation /inspiratory oxygen fraction; ARDS: acute respiratory distress syndrome; Brescia-COVID19: respiratory severity scale Brescia-COVID-19; CT: computed tomography scan; IL-6: Interleukin 6; DRVr: ritonavir-boosted darunavir

**Appendix Figure 1.**
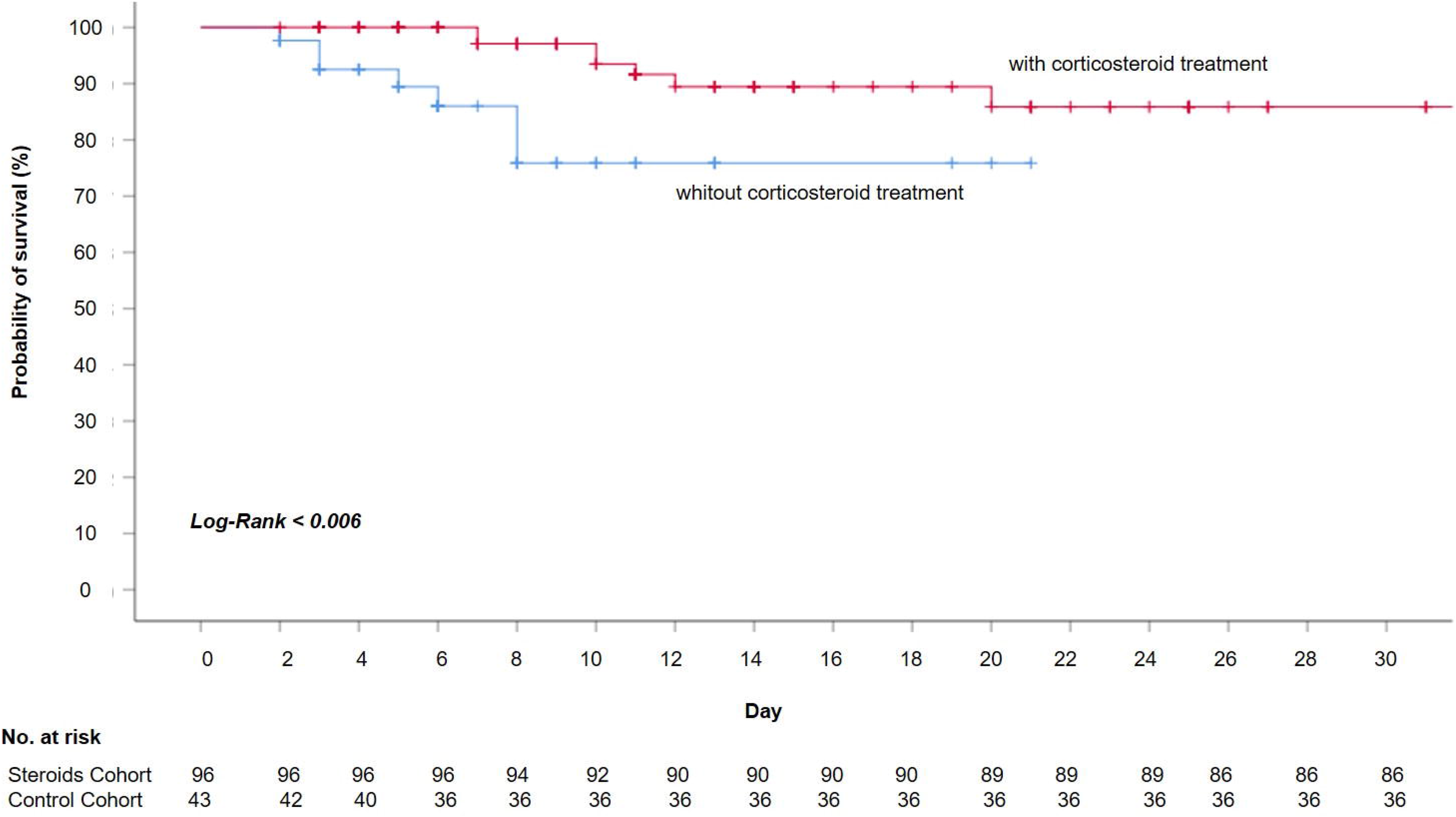
Probability of survival from D0 to hospital discharge of patients with SARS-COV-2 infection, according steroid exposure (Mild ARDS).

**Appendix Figure 2.**
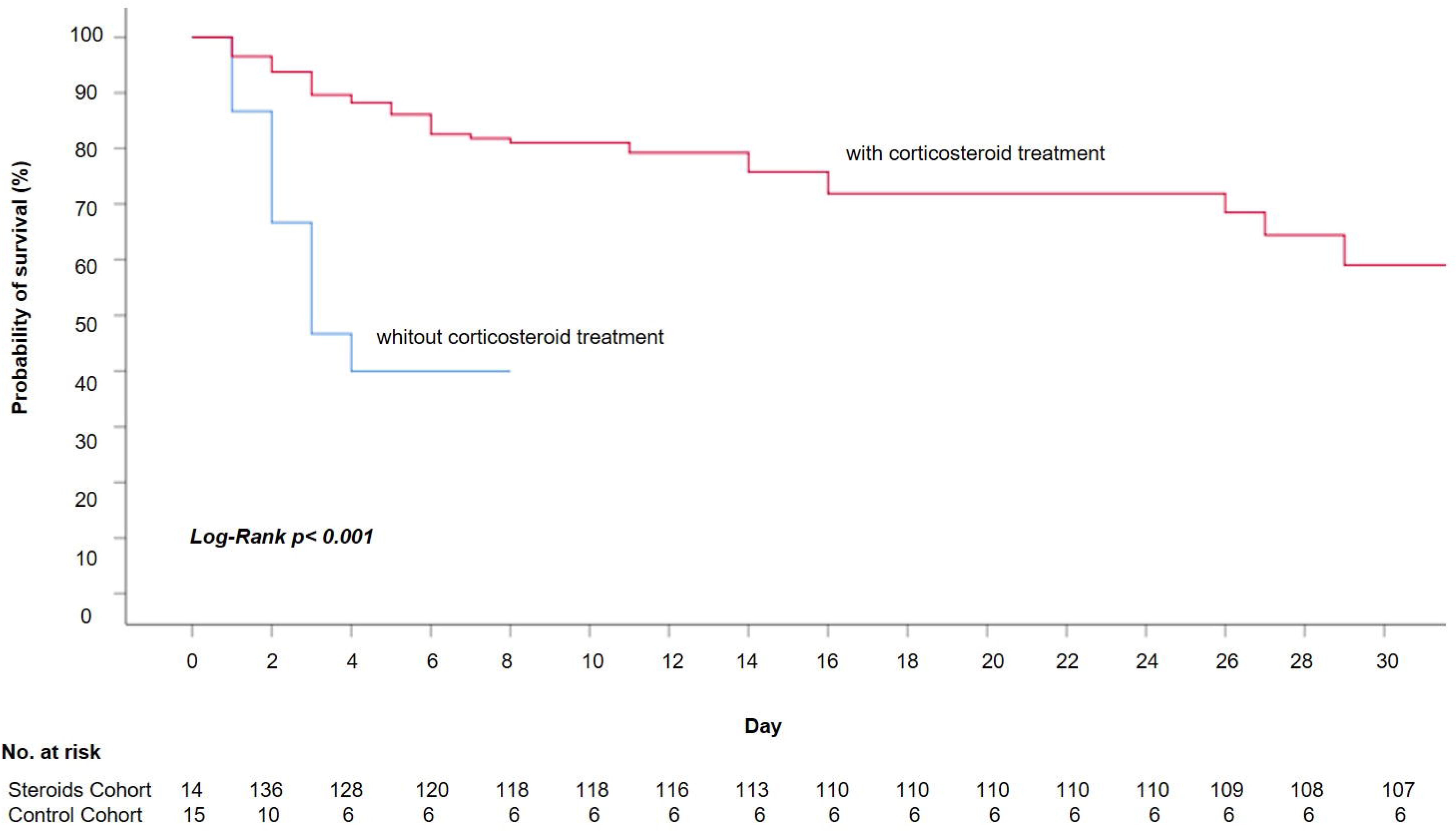
Probability of survival from D0 to hospital discharge of patients with SARS-COV-2 infection, according to steroid exposure (Moderate-severe ARDS).

